# Multiparametric MRI of Early Tumor Response to Immune Checkpoint Blockade in Metastatic Melanoma

**DOI:** 10.1101/2021.05.13.21257127

**Authors:** Doreen Lau, Mary A. McLean, Andrew N. Priest, Andrew B. Gill, Francis Scott, Ilse Patterson, Bruno Carmo, Frank Riemer, Joshua D. Kaggie, Amy Frary, Doreen Milne, Catherine Booth, Arthur Lewis, Michal Sulikowski, Lee Brown, Jean-Martin Lapointe, Luigi Aloj, Martin J. Graves, Kevin M. Brindle, Pippa G. Corrie, Ferdia A. Gallagher

## Abstract

**Background:** Immune checkpoint inhibitors are now standard of care treatment for many cancers. Treatment failure in metastatic melanoma is often due to tumor heterogeneity not easily captured by conventional CT or tumor biopsy. The aim of this prospective study was to investigate early microstructural and functional changes within melanoma metastases following immune checkpoint blockade using multiparametric MRI.

**Methods:** Fifteen treatment-naïve metastatic melanoma patients (total 27 measurable target lesions) were imaged at baseline, and following three weeks and twelve weeks of treatment on immune checkpoint inhibitors using T_2_-weighted imaging, diffusion kurtosis imaging and dynamic contrast-enhanced MRI. Treatment timepoint changes in tumor cellularity, vascularity and heterogeneity within individual metastases were evaluated and correlated to the clinical outcome within each patient.

**Results:** Differential tumor growth kinetics in response to immune checkpoint blockade were measured in individual metastases within the same patient. Early detection of tumor cell death or cell loss measured by a significant increase in the apparent diffusivity D_app_ (*p* < 0.05) was observed in both responding and pseudoprogressive lesions after three week of treatment. Tumor heterogeneity (apparent kurtosis K_app_) was consistently higher in the pseudoprogressive and true progressive lesions, compared to the responding lesions throughout the first twelve weeks of treatment. These preceded tumor regression and significant tumor vascularity changes (K^trans^, v_e_ and v_p_) detected after twelve weeks of immunotherapy (*p* < 0.05).

**Conclusions:** Multiparametric MRI demonstrated potential for early detection of successful response to immunotherapy in metastatic melanoma.

## Background

Immune checkpoint inhibitors targeting the cytotoxic T-lymphocyte antigen-4 (CTLA-4), programmed cell death receptor-1 (PD-1) and programmed cell death receptor-1 ligand (PD-L1) are improving outcomes for increasing numbers of patients with solid cancers^1^. These drugs are now the standard of care for treating many cancers including metastatic melanoma^2^. International trials testing PD-1 antibodies in metastatic melanoma reported response rates of up to 45%^3–6^. Although durable remissions are achieved in some patients, only a minority responded, while all treated patients are at risk of immune-mediated toxicity that can be both life-changing and life-threatening^3,7^. In clinical practice, standard CT and MRI imaging are used for evaluation of treatment response, usually undertaken at twelve weekly intervals. Assessment of response in the early stages can be difficult and can be confounded by possible pseudoprogression, characterized by enlargement of target measurable metastases followed by subsequent regression over time. Biomarkers that could aid clinical decision-making in the first few months of treatment are currently lacking^8^.

Biomarkers derived from whole blood sampling and tumor biopsy do not reflect the spatiotemporal dynamics of tumor immune response to checkpoint inhibition due to the marked inter-patient, inter-metastatic and intra-tumoral heterogeneity present in melanoma^9,10^. Pseudoprogression seen in some patients receiving immune checkpoint inhibitors is difficult to distinguish from true tumor progression using size measurements alone on conventional CT^11,12^. Functional imaging techniques have the potential to longitudinally characterize individual tumor response to immunotherapy and could be used in the future to better predict response.

Several approaches have been investigated to date for imaging response to immune checkpoint inhibition. Positron Emission Tomography (PET) with the glucose analog 2-deoxy-2-[^18^F]fluoro-D-glucose (^18^F-FDG) has shown promise for long-term successful response monitoring: a complete metabolic response (CMR) with ^18^F-FDG uptake 1 year after commencing treatment is associated with an excellent progression free survival compared with those patients who do not show CMR^13^. However, it is not known whether ^18^F-FDG PET can detect early response to treatment, as it can be particularly difficult to distinguish tumor metabolism from glucose uptake associated with immune infiltration after the initial introduction of immune checkpoint inhibitors^14^. Although zirconium-89 radiolabeled antibodies targeting CD8, PD-1 and PD-L1 have been developed as tracers for first-in-human trials in experimental medicine studies^15–17^, these radiolabeled approaches are expensive and cannot be easily implemented as routine clinical tools.

Magnetic resonance imaging (MRI) is a widely available clinical imaging tool. The technique is particularly well-suited for longitudinal tracking of early treatment response, as it does not involve exposure to ionizing radiation^18,19^. Dynamic contrast-enhanced MRI (DCE-MRI) measures properties of tissue vasculature^20^ and is increasingly used in the diagnosis, staging and treatment response assessment of many cancers^21^. Pharmacokinetic modelling of the T_1_-weighted contrast enhanced images provides quantitative measurements of tissue perfusion and vascular permeability^22^. DCE-MRI has been shown to detect tumor perfusion or vascular permeability as a surrogate biomarker of early tumor immune rejection in preclinical models of adoptive T-cell therapy^23,24^, and has been shown to distinguish pseudoprogression from true tumor progression in patients with previously irradiated melanoma brain metastases after three cycles of ipilimumab^25^. Diffusion-weighted imaging is a complementary approach based on the molecular movement of water in tissues which has been widely used for probing changes in cell density due to tumor cell death that occur following successful treatment in cancer^19,26^. An advanced diffusion-weighted imaging approach termed diffusion kurtosis imaging (DKI) has been shown to detect tumor cellularity and heterogeneity in many cancer types based on the non-Gaussian movement of water within the heterogeneous tumor microenvironment^27,28^. Here, we have used a multiparametric imaging approach combining morphological volumetric measurements with DCE and DKI to phenotype the microstructural and functional changes that occur in melanoma metastases before, during and after treatment with immune checkpoint inhibitors.

In this prospective study, early changes in growth kinetics, cellularity, heterogeneity and vascularity of the tumor microenvironment following immune checkpoint blockade between patients and between inter-metastatic lesions were evaluated using multiparametric MRI. Metastatic melanoma offers a paradigm model to test the feasibility of these imaging methods in patients undergoing cancer immunotherapy.

## Methods

### Study design

MelResist is a prospective study approved by the local institutional review board and research ethics committee (11/NE/0312) and managed within the Cambridge Clinical Trials Unit, Cambridge University Hospitals NHS Foundation Trust, Cambridge, UK. Patients were recruited for multiparametric MRI (mpMRI) as part of the MelResist study which evaluated response and resistance biomarkers in metastatic melanoma patients undergoing systemic therapy. Written informed consent was obtained from all patients before enrolment. Patient eligibility criteria for undertaking MRI included: (a) clinical diagnosis of unresectable and previously untreated metastatic melanoma (American Joint Committee on Cancer Stage IV); (b) a treatment plan to commence standard immune checkpoint inhibitors as first line therapy for metastatic melanoma; (c) Eastern Cooperative Oncology Group (ECOG) performance status score of 0 or 1, and life expectancy of twelve weeks or greater; (d) measurable disease on baseline CT (tumor diameter > 1 cm); (e) availability of recent excised or biopsied tissue samples from metastatic tumors for histopathological confirmation; (f) known BRAF V600 mutation status; and (g) no contraindication to MRI.

Enrolled patients received one of the following regimens: (a) anti-PD-1 monotherapy, 2 mg/kg or 200 mg flat dose of pembrolizumab (Keytruda®) every three weeks; or 3 mg/kg or 240 mg of nivolumab (Opdivo®) every two weeks, or 480 mg every four weeks; (b) combined anti-CTLA-4 and anti-PD-1 therapy, 3 mg/kg of ipilimumab (Yervoy®) plus 1 mg/kg of nivolumab (Opdivo®) every three weeks for 4 cycles followed by nivolumab 240 mg every two weeks or 480 mg every four weeks. All treatments were administered by intravenous infusion. Treatment continued until disease progression (as defined by the three-monthly restaging CT scans), development of unacceptable adverse side effects such as autoimmune disorders, or patient withdrawal of consent. A schematic diagram for the mpMRI study flowchart and the clinical characteristics of the study participants are as shown in **Figure 1** and **Table 1**. Further details on the patient demographics can be found in supplemental **Table S1**.

**Table 1.** Clinical characteristics of study participants.

| Characteristics |  |
| --- | --- |
| No. of patients | 15 |
| Age (median age, range) | 65.4 (69, 48 – 76) |
| Gender | 10 M, 5 F |
| AJCC Stage | IV |
| <u>ECOG performance status</u> |  |
| 0 | 9 |
| 1 | 6 |
| <u>BRAF status</u> |  |
| *BRAF mutant* | 3 |
| *BRAF WT* | 12 |
| <u>Serum LDH (IU/mL) at baseline</u> |  |
| *Normal (< 250)* | 10 |
| *Elevated (> 250)* | 5 |
| <u>Neutrophils-to-lymphocyte ratio at baseline</u> |  |
| *Normal (< 5)* | 12 |
| *High (> 5)* | 3 |
| <u>Immunotherapy</u> |  |
| *Pembrolizumab* | 6 (40.0 %) |
| *Nivolumab* | 2 (13.3 %) |
| *Combined Ipilimumab and Nivolumab* | 7 (46.7 %) |
| <u>CT Evaluation at 12<sup>th</sup> week</u> |  |
| *Partial Response* | 6 (40.0 %) |
| *Mixed Response* | 4 (26.7 %) |
| *Progressive Disease* | 5 (33.3 %) |
| <u>CT Evaluation at 12<sup>th</sup> month</u> |  |
| *Partial Response* | 7 (46.7 %) |
| *Progressive Disease* | 8 (53.3 %) |
| <u>Anatomical site selected for MRI Imaging</u> |  |
| *Head & Neck* | 2 |
| *Chest* | 1 |
| *Abdomen & Pelvis* | 7 |
| *Subcutaneous* | 4 |
| *Limbs* | 1 |

*All patients imaged were histologically confirmed as AJCC Stage IV melanoma.*
*AJCC = American Joint Committee on Cancer staging (7<sup>th</sup> edition);*
*ECOG = Eastern Cooperative Oncology Group; LDH = Lactate Dehydrogenase*

**Figure 1.**
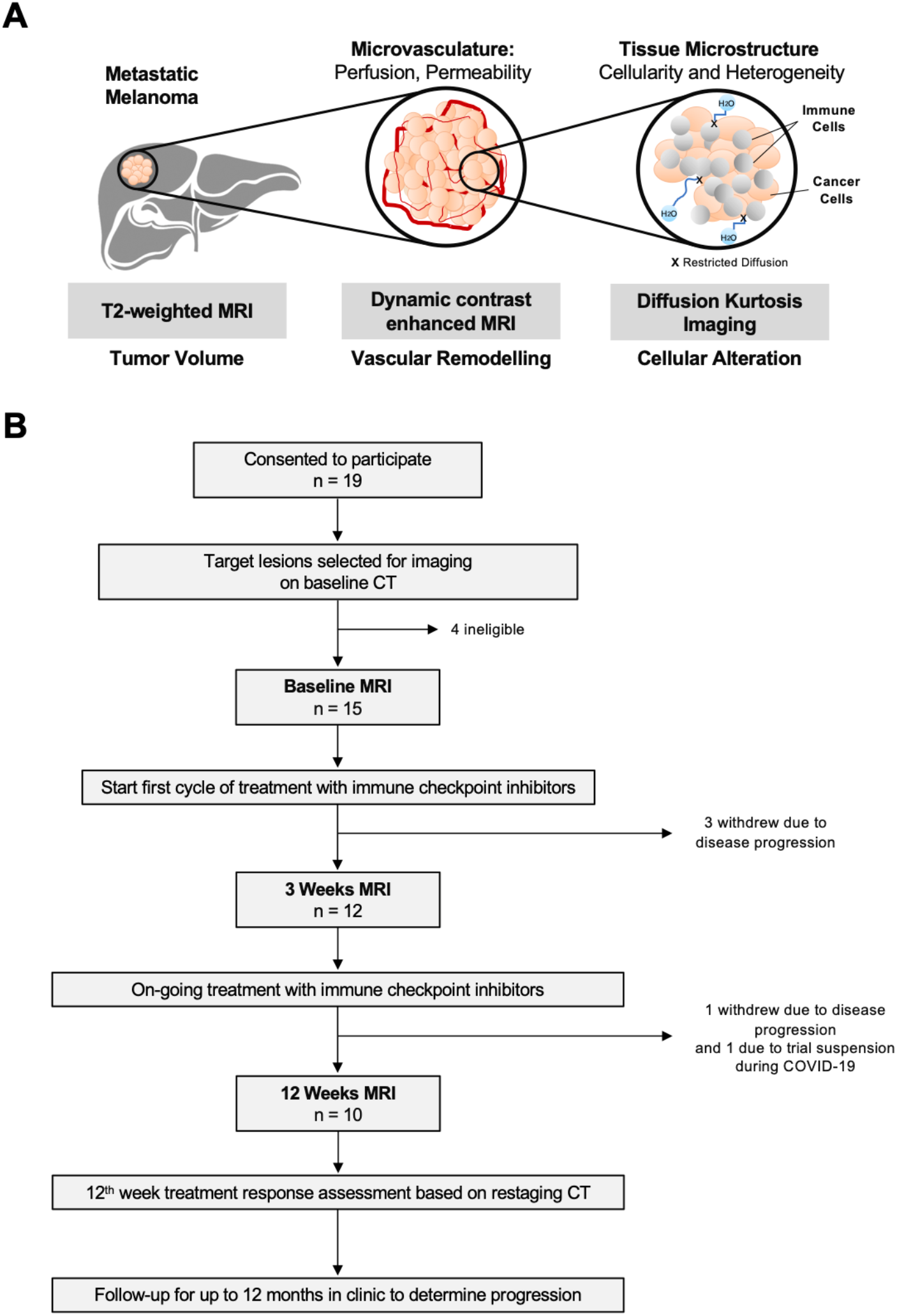
A mpMRI approach for longitudinal tracking of biological changes within tumors in response to immune checkpoint blockade. (**A**) Schematic diagram of the mpMRI approaches used in this study for monitoring tumor response to immune checkpoint blockade. (**B**) Study flowchart for the melanoma immunotherapy trial (MelResist).

### MR imaging

All patients underwent proton (^1^H) MRI on a 3.0 Tesla system (Discovery MR750, GE Healthcare, Waukesha, WI) using a 32-channel phased-array coil with respiratory-gating or multiple breath-holds used to reduce motion artifacts during image acquisition for lesions in the abdomen. The mpMRI protocol included multiplanar T_2_-weighted single-shot fast spin-echo anatomical imaging, diffusion kurtosis imaging (DKI) of tumor cellularity and heterogeneity, and dynamic contrast-enhanced MRI (DCE-MRI) of tumor perfusion or vascular permeability. Imaging was conducted at three timepoints: within one week prior to starting treatment with immune checkpoint inhibitors (<u>Baseline</u>); three weeks after the first infusion (<u>3-Weeks</u>) and twelve weeks after the start of treatment (<u>12-Weeks</u>) coinciding approximately with the first standard restaging CT response assessment. Further details on the imaging acquisition, image processing and analysis can be found in the **Supplemental Material** and **Table S2**.

### Classification of target melanoma metastases and measurement of response

Conventional objective response of the target metastases was determined by measuring the best treatment outcome at the 12^th^ week restaging CT, and follow-up at 12 months. Metastases with more than 30% decrease in volume on the 12-Weeks MRI were classified as responding, metastases with more than 20% increase in volume were identified as true progression, while metastases with more than 20% increase in volume at the 3-Weeks MRI but which subsequently decreased in >30% volume on the 12-Weeks scans were classified as pseudoprogression.

### Statistical analysis

Statistical analysis was performed in GraphPad Prism software version 8 (La Jolla, CA, USA). All values were expressed as median and interquartile range to account for sample size differences between groups. Normality was assessed using the Shapiro-Wilk test. Changes in individual lesion mpMRI biomarkers over the treatment timepoints were evaluated using either paired t-test for normally distributed data or Wilcoxon matched-pair signed-rank test for data with non-parametric distribution. Differences between the subgroups of responding, pseudoprogressive and true progressive lesions were evaluated using one-way ANOVA for normally distributed data, or the Kruskal-Wallis test with Dunn’s multiple comparison for non-parametric testing. Spearman’s correlation analysis was used for evaluating any relationship between the mpMRI biomarkers across treatment timepoints. A value of *p* < 0.05 was considered as statistically significant.

## Results

### Clinical characteristics

Fifteen treatment-naïve patients (10 males, 5 females; median age 65 years) were imaged with mpMRI over the first twelve weeks of immunotherapy. 10 patients completed MRI at all three imaging timepoints (Baseline, 3-Weeks and 12-Weeks); 5 patients were scanned at Baseline and/or 3-Weeks before withdrawal from the trial due to clinical reasons such as early disease progression or clinical deterioration incompatible with continuing on the study. An additional 4 patients were enrolled on the study but were deemed as ineligible for the prospective trial due to insufficient time for scheduling of imaging scans before the start of treatment (within a week) or target lesions that were too small (less than 1 cm in largest diameter) for multiple timepoint imaging and follow-up treatment response assessment. 53% of the patients received PD-1 monotherapy, while 47% of the patients were treated with combined CTLA-4 and PD-1 therapy.

### Differential response to immune checkpoint blockade

At the 12^th^ week restaging CT evaluation, 6 patients demonstrated partial response to immune checkpoint inhibitors and 5 patients showed disease progression. There were 9 patients who had more than 1 target lesion imaged. Of these, 4 patients demonstrated a mixed response between the individual metastases. 3 out of these 4 patients subsequently progressed six months after the start of treatment, whereas 1 patient continued to respond to treatment (**Table 1**). Further details on patient demographics can be found in supplemental **Table S1**.

The mpMRI images for a total number of 27 enhancing target melanoma metastases that were first identified as more than 1 cm in diameter on staging CT were analyzed. In this study, a total of 13 responding, 4 pseudoprogressive and 10 true progressive metastases were identified. There were no lesions which showed a 30% decrease in volume at the 3-Weeks MRI which subsequently increased in volume at the 12-Weeks MRI and restaging CT.

T_2_-MRI volumetric analysis showed differential inter-patient and inter-metastatic response to immune checkpoint blockade. Within the cohort of patients in our study, inter-metastatic differences in the individual tumor growth kinetics were particularly evident in patients undergoing PD-1 monotherapy, as compared to patients receiving combined CTLA-4 and PD-1 treatment where response was almost immediate at the 3-Weeks MRI (**Figure 2A**). Interestingly, increasing T_2_ hyperintensity or inflammatory changes were detected within all 4 enlarged pseudoprogressive tumors at 3-Weeks, which resolved at 12-Weeks with a corresponding reduction in tumor volume (**Figure 2B** and supplemental **Figure 3**).

**Figure 2.**
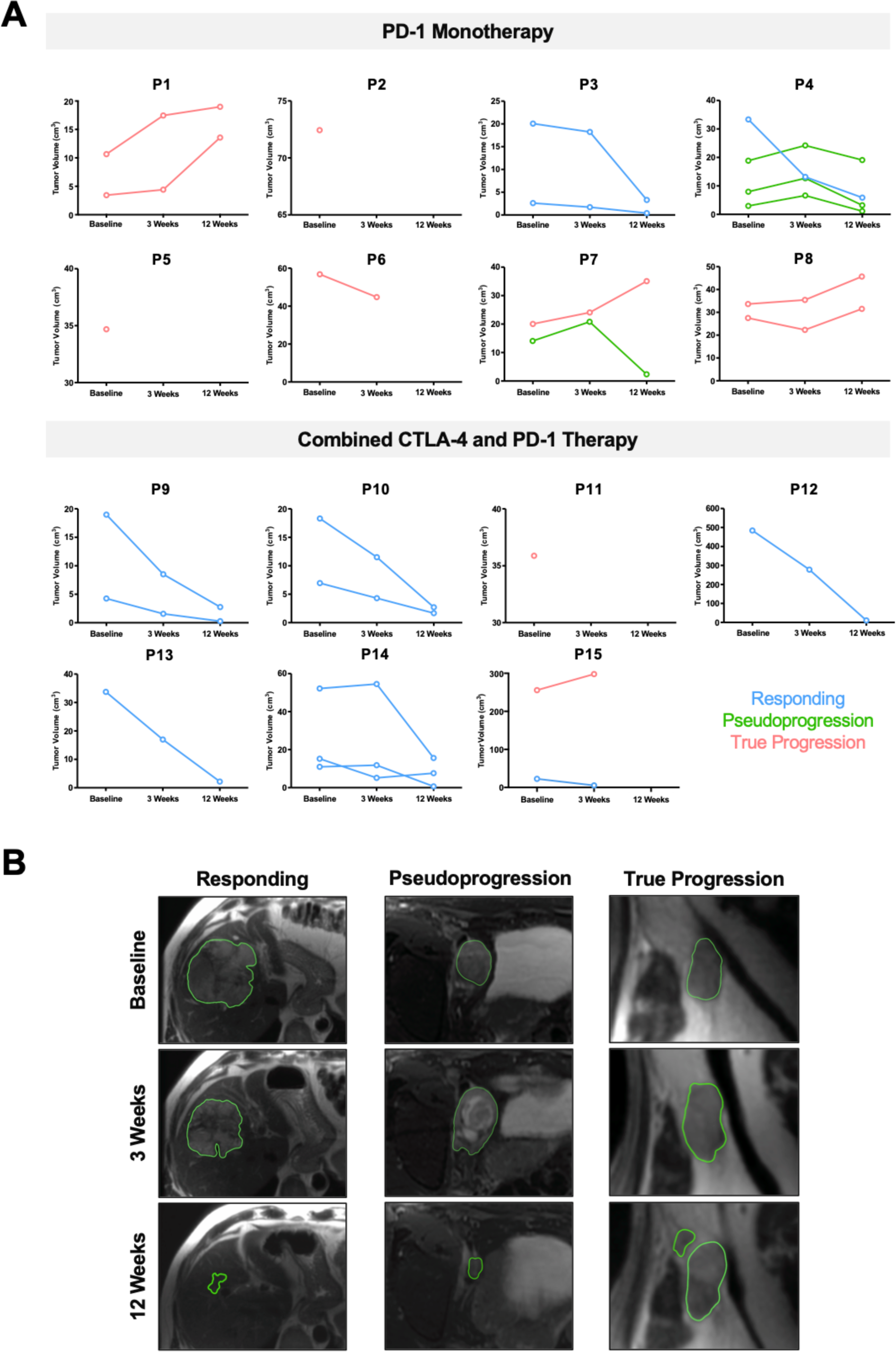
Inter-patient and inter-metastatic heterogeneity in response to immune checkpoint blockade. (**A**) Differential tumor growth kinetics in patients receiving PD-1 monotherapy compared to combined CTLA-4 and PD-1 treatment. Individual tumor volumes were measured on T_2_-weighted MRI. Categorization of tumors into three subgroups (Responding, Pseudoprogression and True Progression) were based on assessment at 12 months. (**B**) Representative T_2_-weighted images from three patients with the classic features of responding, pseudoprogressive and true progressive lesions. Note the T2 hyperintensity in keeping with inflammation in the pseudoprogressive lesion at 3-Weeks.

### Tumor cell death and changes in heterogeneity in response to treatment

**Figure 4** and supplemental **Figure S1** show the changes in tumor cellularity and heterogeneity measured on DKI. No significant difference in the average apparent diffusivity (D_app_) for each patient, as a measure of tumor cell density, was detected between the responding and non-responding patients at Baseline (**Figure 4A**; median D_app_ of 1.44 for responding patients versus 1.33 for non-responding patients, *p* = 0.62). There was a significant increase in the average D_app_ of imaged target metastases for each patient representing reduced tumor cellularity (*p* < 0.05) in the responding patients at 3-Weeks (median D_app_ 1.65; interquartile range (IQR) 1.59–1.77) compared to Baseline (1.44; IQR 1.26 – 1.63), with a further significant increase at 12-Weeks (2.01; IQR 1.60 – 2.22). In contrast, there was no significant change in D_app_ in the tumors of non-responders over the twelve weeks of treatment (**Figure 4B**). Further analysis based on classification of individual metastases from all patients into the three subgroups of “Responding”, “Pseudoprogression” and “True Progression”, showed a significantly lower D_app_, reflecting higher tumor cell density at Baseline in the pseudoprogressive lesions (median 1.17; interquartile range (IQR) 1.02 – 1.20), as compared to the responding (median 1.48; IQR 1.44 – 1.68; *p* < 0.001) and true progressive lesions (median 1.44; IQR 1.15 – 1.82; *p* < 0.05). Individual tumors responded differently to treatment: most of the responding and pseudoprogressive lesions exhibited a significant percentage increase in D_app_ at the 3-Weeks MRI relative to baseline, indicating lower cellularity in most responding lesions (median increase in D_app_ by 8.9%; IQR 2.3 – 27.6%; *p* < 0.05) and pseudoprogressive lesions (median increase by 48.0%; IQR 45.2 – 63.1%; *p* < 0.05). A further increase in D_app_ was detected within the tumor microenvironment in most of the metastases responding at 12-Weeks (31.7%; IQR 1.9 – 45.5%; *p* < 0.05). However, one lesion demonstrated higher cellularity (increase in D_app_) despite a reduction of tumor volume over twelve weeks of treatment: interestingly, this lesion subsequently increased in size at the 6^th^ month restaging CT and was verified to be a pseudoprogressive lesion over a longer timeframe. Higher cellularity was also detected on average in the pseudoprogressive lesions at 12-Weeks compared to the responding lesions, despite a reduction in tumor volume, which may reflect a later phase of immune infiltration and tumor cell killing in these metastases (**Figure 4** and **Figure 5**). The D_app_ values measured from the experimental DKI images correlated to the apparent diffusion coefficient (ADC) values obtained on clinical DWI (supplemental **Figure S2**).

**Figure 3.**
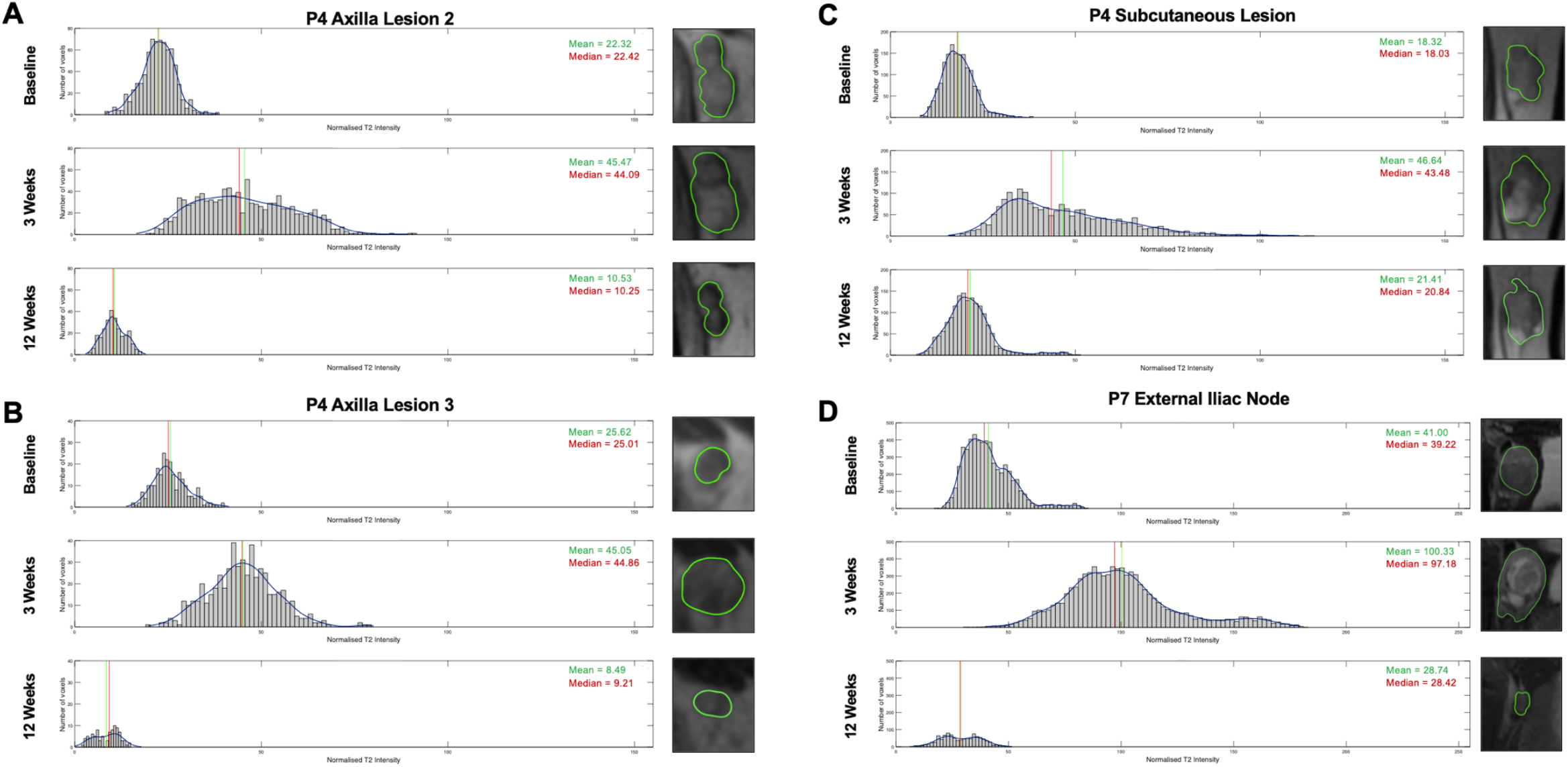
Histogram analysis of the T_2_ intensity values of all 4 pseudoprogressive lesions. These included: the (**A**) axilla lesion 2, (**B**) axilla lesion 3, (**C**) subcutaneous lesion of Patient P4; and the (**D**) external iliac node of Patient P7.

**Figure 4.**
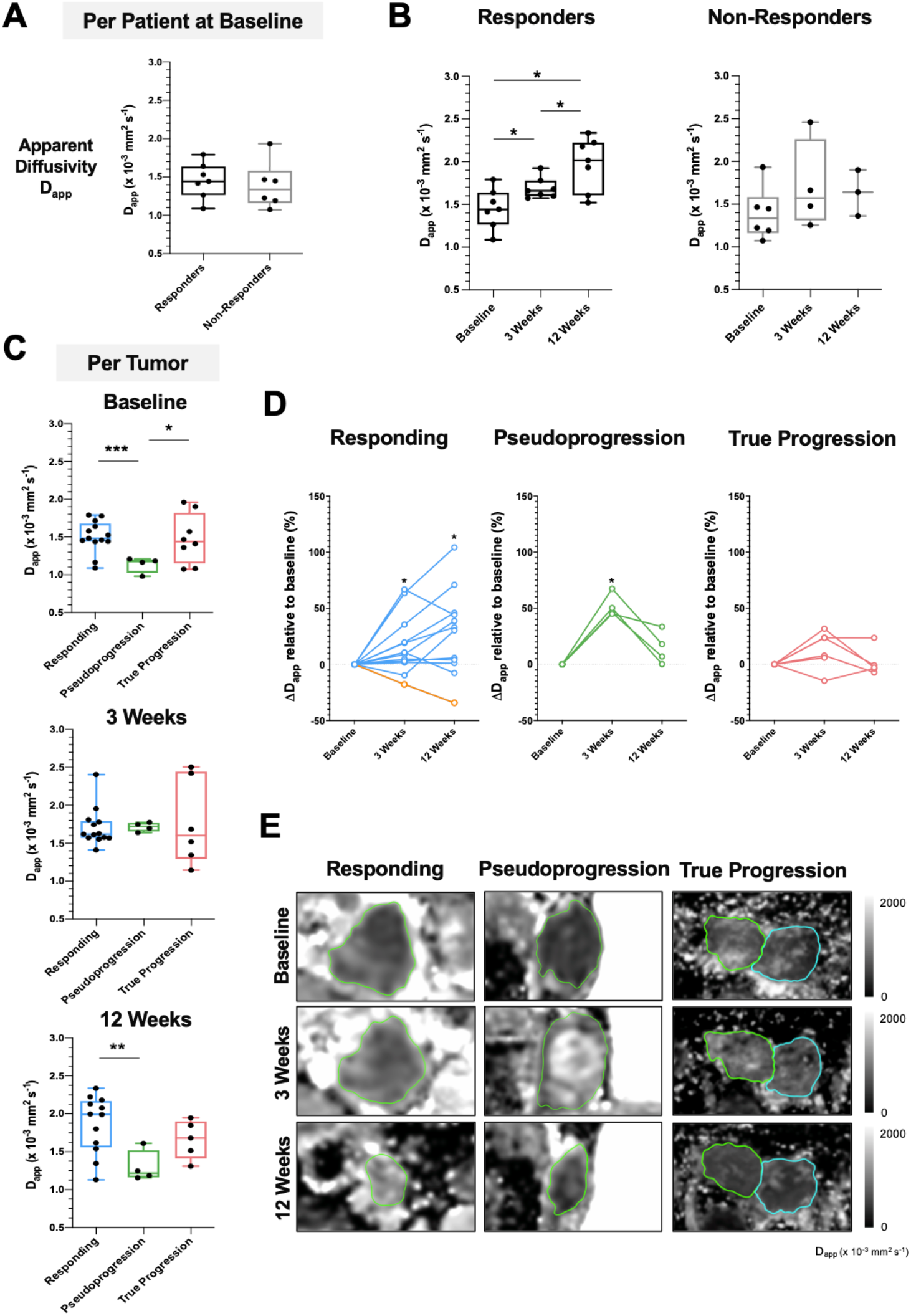
Early detection of tumor cell death using DKI. (**A**) Comparison of apparent diffusivity (D_app_) as a measure of tumor cell density between responders and non-responders at baseline before the start of treatment. (**B**) Changes in average tumor D_app_ on a per patient basis over the course of treatment, divided according to overall response. (**C**) Response of individual lesions classified into three subgroups (Responding, Pseudoprogression, and True Progression) showing the differences in tumor cellularity at baseline. (**D**) Percentage change in D_app_ relative to baseline in individual lesions from the three subgroups. (**E**) Representative D_app_ images from three lesions categorized as Responding, Pseudoprogression, and True Progression respectively based on the 12^th^ month restaging CT. Data are presented as median and interquartile range. Normality was assessed using the Shapiro-Wilk test. Mann-Whitney test was performed to assess differences between two independent lesion subgroups; Kruskal-Wallis test with *post hoc* Dunn’s multiple comparison analysis was performed to test for differences between three independent lesion subgroups; * *p* < 0.05; *** *p* < 0.001. Yellow line in (D) indicates the percentage change in D_app_ for patient P4. Analysis of apparent kurtosis (K_app_) as a measure of tumor heterogeneity, detected concurrently using DKI, is found in supplemental **Figure S1**.

**Figure 5.**
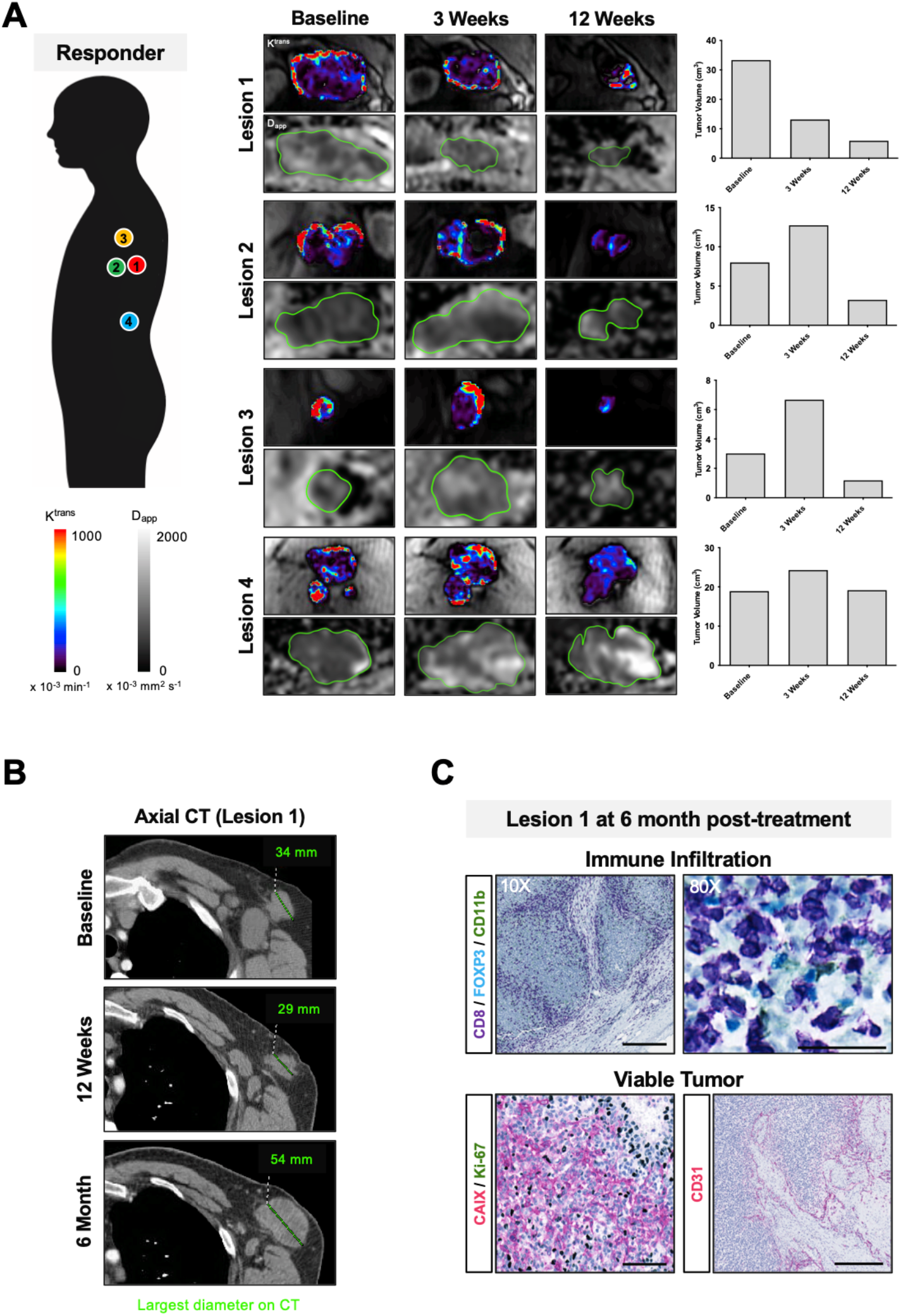
Dynamic changes in intertumoral response to immune checkpoint blockade within a single patient. (**A**) Multiple metastases in a patient on treatment with Nivolumab (PD-1 monotherapy). Intertumoral differences in treatment response, vascular permeability and cellularity were measured in four target lesions on MRI during the first 12 weeks of treatment. Increased cellularity was detected in the responding tumor (Lesion 1) despite a reduction in tumor volume and lower vascularity at 12-Weeks. The responding lesion subsequently progressed at 6^th^ month on treatment and was surgically resected. (**B**) Axial CT of Lesion 1 at baseline, 12^th^ week and 6 months; largest tumor diameter shown in mm. (**C**) Immunohistochemistry of Lesion 1 showed remarkable infiltration of immune cells (CD8) in viable tumor tissues that were highly hypoxic (CAIX), proliferative (Ki67) and vascular (CD31). Scale bars for CD8 immunostained images represent 100 µm (10x magnification) and 50 µm (80x magnification); 100 µm in CAIX/Ki67 dual staining (20x magnification) and CD31 (10x magnification).

No significant difference in microscopic tumor heterogeneity at Baseline (*p* = 0.27) was detected in the tumors of the responders (median K_app_ 0.61; IQR 0.51 – 0.71), compared to non-responding patients (median 0.71; IQR 0.53 – 0.86), as detected by the average apparent kurtosis (K_app_) values from all target lesions for each patient obtained concurrently on DKI (supplemental **Figure S1**). A significant reduction in K_app_ (*p* < 0.05) was detected in the tumors of responding patients at 3-Weeks (median 0.59; IQR 0.51 – 0.65) compared to Baseline (median 0.61; 0.51 – 0.71). Further analysis of the individual lesions showed a trend towards a higher level of tumor heterogeneity, as measured by K_app_, in the pseudoprogressive lesions throughout the first twelve weeks of treatment (Baseline: 0.84; 3-Weeks: 0.64; 12-Weeks: 0.76) compared to the responding lesions (Baseline: 0.59; 3-Weeks: 0.54; 12-Weeks: 0.56); although this did not reach statistical significance, it may reflect underlying immune cell infiltration or cell death over the course of treatment. As with the results for D_app,_ no significant change in K_app_ was detected in the progressing metastases during the first twelve weeks of treatment.

### Tumor vascular remodeling following cell death

**Figure 6** and supplemental **Figure S3** show the changes in tumor vascularity and perfusion during twelve weeks of treatment, as measured by DCE-MRI and contrast kinetic modelling using the extended Tofts model. The average tumor vascular transfer constant (K^trans^) at baseline was higher in the target lesions of the responding patients (median K^trans^ 0.56; IQR 0.23 – 1.37) compared to the non-responders (0.15; IQR 0.11 – 0.44; *p* < 0.05). Similarly, the average fractional extravascular-extracellular volume (v_e_) at Baseline of the target lesions of the responding patients were higher (median v_e_ 0.49; IQR 0.31 – 0.77) compared to the non-responders (0.19; IQR 0.15 – 0.32; *p* < 0.05). A significant reduction in these tumor vascularity metrics (K^trans^, v_e_ and v_p_) was only detected at 12-Weeks compared to Baseline (0.11; IQR 0.05–0.45; *p* < 0.05) but not at 3-Weeks. A gradual increase in tumor vascular metrics was detected in the tumors of non-responding patients over the course of treatment but this was not statistically significant which may reflect the small numbers, particularly at 12-Weeks. Further analysis on the individual lesions showed no significant difference in the vascular transfer constant K^trans^, fractional volume of the extravascular-extracellular space v_e_, or fractional plasma volume v_p,_ between the three subgroups of lesions before the start of treatment. A significant decrease in K^trans^ relative to Baseline was detected in most responding lesions at 12-Weeks (median −66.19%; IQR −92.00 to −46.49%; *p* < 0.01), but not at 3-Weeks (−29.73%; IQR −40.51 to 14.01%; *p* = 0.23). Similarly, v_e_ and v_p_ were also lower in the responding lesions at 12-Weeks (*p* = 0.07 and *p* < 0.01 respectively). A trend towards lower K^trans^ was also detected in most pseudoprogressive lesions at 3-Weeks (median 0.47; IQR 0.18 – 0.60) and 12-Weeks (0.15; IQR 0.11 – 0.32) compared to Baseline (0.52; IQR 0.19 – 0.82), but this was not statistically significant given the small number of pseudoprogressive lesions within the patient cohort.

**Figure 6.**
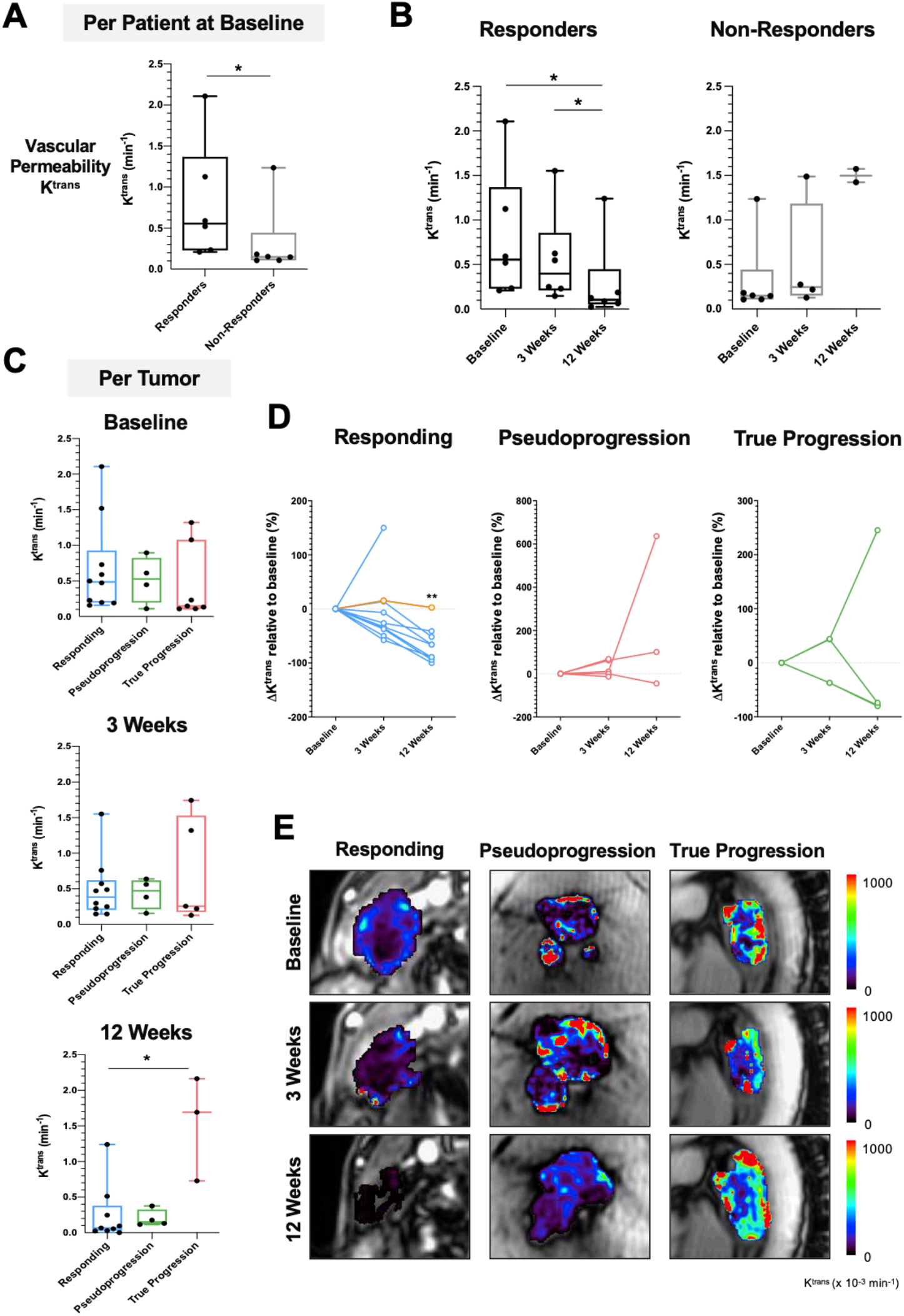
Tumor vasculature remodeling following immune cytotoxic killing and tumor cell death. (**A**) Comparison of vascular permeability K^trans^ between responders and non-responders at baseline before the start of treatment. (**B**) Changes in tumor K^trans^ over the course of treatment. (**C**) Comparison of tumor vascular permeability at baseline between individual lesions from the three subgroups: Responding, Pseudoprogression, and True Progression. (**D**) Percentage change in K^trans^ relative to baseline in individual lesions from the three subgroups. (**E**) Representative K^trans^ images from the three subgroups lesions. * *p* < 0.05; ** *p* < 0.01. Yellow line in (D) indicates the percentage change in K^trans^ for patient P4. Analysis for other DCE-MRI parametric measurements is found in supplemental **Figure S3**.

### Early treatment timepoint changes in tumor cellularity is independent of tumor volume

Spearman’s correlation analysis of the mpMRI biomarkers over the first twelve weeks of treatment in this cohort showed no significant correlation between tumor volume and D_app_ (supplemental **Figure S4**). This implies that the early detection of changes in tumor cellularity from immune cytotoxic killing of tumor cells at the 3-Weeks MRI was independent of changes in tumor volume. However, a positive correlation was found between tumor volume and all metrics of tumor vascularity and perfusion at 12-Weeks (K^trans^, v_e_ and v_p_), suggesting that vascular remodeling may be related to tumor size changes following immune checkpoint blockade.

## Discussion

As immune checkpoint inhibitors become more widely used in routine clinical practice, there is an unmet need for more effective tools to measure successful response to these agents. This is increasingly important as more cancer patients are being offered long term immunotherapy, which is not only costly to health care systems, but also comes with a significant risk of side effects. Tumor heterogeneity is one of the major challenges for effective cancer treatment and manifest as morphological, functional, cellular, metabolic and molecular diversity^29,30^. Clinical tools to image this multilayered tumor heterogeneity and how it changes with immunotherapy, could have a role in differentiating tumor resistance from successful response early in the treatment pathway^31^.

In this study, longitudinal tracking of microstructural and functional changes in metastatic melanoma during the first twelve weeks of treatment was performed using mpMRI. Heterogeneity in response to immune checkpoint blockade was observed in these treatment-naïve tumors. This heterogeneity may be due to inter-patient and inter-metastatic differences in tumor immunogenicity, as some patients may have more delayed response, multiple waves of immune activation or on-going immune evasion and clonal expansion during continuous treatment as exemplified by in patient P4. These changes in the tumor microenvironment may not be identified by standard CT or MRI imaging, as the change in tumor size alone are often insufficient to determine treatment benefit at the early stages of immunotherapy. Therefore, a clinically applicable tool to evaluate immunotherapy is required to guide clinical decision-making^32^.

Treatment response to immune checkpoint blockade was captured longitudinally on mpMRI in this study using three approaches to assess the tumor microenvironment: T_2_-weighted MRI of tissue structure, DKI of cellular density and its microscopic heterogeneity, and DCE-MRI of the tumor vasculature. An interesting and unexpected observation was an increase in median T_2_-weighted signal intensity following three weeks of immunotherapy (one infusion of immunotherapy) in the pseudoprogressive metastases, compared to metastases that responded, or were shown to progress at later timepoints. This is likely to be due to tumor enlargement from significant immune cell infiltration and inflammation, rather than tumor proliferation. T_2_-weighted MRI represents a very simple routine clinical tool which may be able to discriminate pseudoprogression from true progression if this initial observation could be confirmed in larger studies. Quantitative analysis of these signal intensity changes e.g. using T_2_ mapping may be useful in future trials for more detailed characterization of these microstructural changes^33^.

Changes in cell density were measured using apparent diffusivity (D_app_) on DKI to detect either cytotoxic T-cell killing or increased cell density from immune infiltration or tumor proliferation. Cell loss measured as an increase in the median D_app_ was detected in both responding and pseudoprogressive lesions as early as three weeks after the start of treatment compared to Baseline. Further reductions in cell density within the responding lesions were detected at the 12-Weeks MRI. An increase in ADC measured on diffusion-weighted imaging (equivalent to the D_app_ measured here) has also been reported in previously treated ocular melanoma responding to immunostimulatory adenoviral CD40L gene therapy: ≥ 1-fold change in ADC at week 5 following treatment was a better predictor of objective survival than metabolic changes on ^18^F-FDG PET and tumor size changes on MRI^34^. Interestingly, in our study, an increase in cell density (or lower D_app_) was detected in the pseudoprogressive lesions at 12-Weeks compared to the responding lesions despite reduced tumor volumes measured on the 12-Weeks MRI and standard restaging CT. No significant correlation between D_app_ and tumor volume was detected across the imaging timepoints, which implies that the estimation cell density based on water diffusion within the tumor microenvironment is independent of tumor volume and is therefore an important additional metric to measure. A higher degree of tumor heterogeneity, as assessed by an increase in apparent kurtosis K_app_, was detected in the pseudoprogressive lesions throughout the MRI imaging timepoints, compared to the responding lesions. This supports the hypothesis that there is underlying cellular alteration with different phases of immune activation and proliferation in the pseudoprogressive lesions over the course of treatment. Although there was a higher K_app_ in the true progressive lesions at all imaging timepoints compared to the responding lesions, the feasibility of using DKI alone to differentiate true progression from pseudoprogression could not be established as the number of non-responders with complete MRI scans are limited in the metastatic disease setting due to early disease progression and withdrawal from the trial. Nevertheless, greater tumor heterogeneity at baseline (entropy, dissimilarity and contrast texture features) measured on CT radiomics has been previously reported in non-responders to PD-1 monotherapy^35^.

The tumor vasculature plays a significant role in regulating tumor homeostasis, metastasis and immune trafficking^36^. The vascular networks in malignant tumors are typically disorganized with immature, tortuous, and leaky blood vessels that are hyperpermeable to intravascular contrast agents. DCE-MRI showed a gradual decrease in: the tumor vascular permeability K^trans^; the extravascular-extracellular space v_e_; and the plasma volume fraction v_p_ within the responding lesions. This was more prominent at the 12-Weeks MRI when a reduction in tumor burden was detected, suggesting that tumor vasculature remodeling and shutdown may have occurred following cell death caused by cytotoxic T-cell killing. This contrasts with the effects of anti-angiogenic treatments in human melanoma xenografts whereby the treatments are more directed towards the vascular network and are generally not cytotoxic. Tumor vasculature remodeling i.e. lower K^trans^ often precedes cell death and reduction in tumor burden, with no significant change in cell density measurements, v_e_ or ADC^37^. Despite the small number of pseudoprogressive lesions available for analysis in our study, lower K^trans^, v_e_ and v_p_ with a corresponding reduction in tumor volume as a result of cell death was detectable in most pseudoprogressive lesions at 12-Weeks. The pseudoprogressive lesions in general demonstrated lower vascular permeability and perfusion compared to the true progressive lesions at 12-Weeks. Our findings are in concordance with a previous study assessing DCE-MRI melanoma brain metastases study in which lower v_p_ was detected in previously irradiated pseudoprogressive lesions compared to true progressive lesions after three cycles of ipilimumab^25^. This suggested that DCE-MRI may have utility in distinguishing true progressive lesions from treatment-responsive lesions, but at a later timepoint compared to diffusion measurements. Interestingly, higher K^trans^ and v_e_ were detected at Baseline in all imaged tumors of most patients who were responders to immune checkpoint inhibitors compared to the non-responders. One explanation could be that the differences in tumor vasculature between tumors may play a role in determining immune trafficking and subsequent immune eradication of tumor cells^36^. Ideally, this could be explored by tissue sampling of multiple lesions both before and during therapy, but this is not practical in the metastatic disease setting clinically and further pre-clinical research is required.

Our study presented several strengths and limitations. This is the first prospective MRI study to serially track cellular and functional changes in melanoma metastases during immune checkpoint blockade. As all melanoma metastases analyzed in this study were previously untreated and unresectable tumors, the treatment timepoint changes measured on mpMRI were directly related to immunotherapeutic effects on individual lesions. Partial volume effects on image measurements were minimal as several patients in our trial presented large metastases at baseline. A stringent criterion for imaging and analysis was maintained to include only patients with measurable disease so that the biological changes were trackable over twelve weeks of immunotherapy. Nevertheless, our study is limited by the small sample size, which restricts the scope for wider interpretation of the results and evaluation of the imaging biomarkers for their predictive values. Future multicenter trials are required to test and validate these imaging biomarkers in a larger patient cohort, with the aim of integrating these imaging methods into immunotherapy trials and routine clinical management. Our study is further limited by the lack of radiologic-pathological correlation, as relatively few metastases are readily accessible to biopsy. This difficulty in obtaining tumor tissues from metastatic sites further highlights the strengths of non-invasive imaging as a surrogate for pathology as changes in tumor growth kinetics, cell density, heterogeneity and vascularity within individual tumors could be longitudinally tracked over the course of treatment.

As part of this prospective study, we have demonstrated marked intralesional, inter-metastatic and inter-patient heterogeneity in melanoma over the first twelve weeks of immunotherapy. After only three weeks of treatment i.e. one infusion of immunotherapy, a decrease in cellularity as measured on DKI, could distinguish responding patients from non-responders, as well as individual responding and pseudoprogressing tumors from true progressing ones. An interesting finding was an increase in normalized T_2_-weighted signal and its distribution in the pseudoprogressing lesions compared to the progressing lesions after three weeks treatment. Therefore, combining conventional T_2_-weighted and DKI at three weeks after starting immunotherapy could be used to identify pseudoprogression during the early stages of treatment. Although there was higher tumor vascular permeability and perfusion at Baseline in the responding patients on DCE-MRI compared to non-responders, changes in K^trans^ could not be used to distinguish responding and pseudoprogressing lesions until after twelve weeks of treatment showing that measurable vascular changes occur later than changes in cellularity.

In conclusion, mpMRI has shown potential for early assessment of response to immunotherapy in metastatic melanoma patients. Early changes in tumor cellularity measured on DKI following three weeks after starting treatment could be used to detect responding and pseudoprogressive melanoma metastases before a change in tumor volume and vascular permeability. This work could have important implications not only for monitoring treatment of metastatic melanoma, but also the increasing number of solid cancers treated with immunotherapy.

## Data Availability

All data relevant to the study are included in the article or uploaded as supplementary information. Data are available upon reasonable request. Data may be obtained from a third party and are not publicly available. Please contact the corresponding authors.

## Declarations

### Ethics approval and consent to participate

The protocol for this study was reviewed and approved by the local institutional review board and research ethics committee in UK (11/NE/0312). Written informed consent was obtained from all patients before enrolment.

### Consent for publication

Consent for publication has been obtained from all authors and study participants.

### Availability of data and material

Data are available upon reasonable request. Please contact the corresponding author Doreen Lau

### Authors’ Contribution

Conceptualization: DL, FAG, PGC, MAM, MJG and KMB. Data curation and formal analysis: DL, FAG, PGC, MAM, ANP, ABG and FS. Investigation: DL, FAG, PGC, MAM and ANP. Methodology: DL, MAM, ANP, ABG, IP, BC, FR, JDK, MJG, MS and LB. Resources: AF, DM, CB, AL, MS, LB, JML and LA. Writing – original draft: DL, FAG and PGC. Writing – review and editing: DL, MAM, ANP, ABG, FAG and PGC.

## Acknowledgements

This project was supported by Cancer Research UK (CRUK; C19212/A16628, C19212/A911376), the CRUK Cambridge Centre (C9685/A25177), a Cambridge Commonwealth, European and International Trust PhD Scholarship, a European Institute of Innovation and Technology (EIT) health grant for Innovation for Personalised Cancer Medicine, the CRUK & Engineering and Physical Sciences Research Council (EPSRC) Cancer Imaging Centre in Cambridge and Manchester (C197/A16465), Cancer Core Europe, the European Institute of Innovation and Technology (EIT) and Addenbrooke’s Charitable Trust. This research was also supported by the National Institute for Health Research (NIHR) Cambridge Biomedical Research Centre (RG85317). The views expressed are those of the author(s) and not necessarily those of the NIHR or the Department of Health and Social Care. Special thanks to Prof. Edwin Chilver and Prof. Klaus Okkenhaug for the PhD co-supervision of Doreen Lau; our research trial patients for supporting this study; Cambridge Cancer Trials Centre clinical trial coordinator David Bruce and Addenbrooke’s Hospital Human Research Tissue Bank for assistance on FFPE samples; Stephanie Heasman, Sofia Koch, Christopher Bagnall, Hannah Hibbs and Elizabeth Henley from AstraZeneca UK for the logistics support in pathology; and the Magnetic Resonance & Spectroscopy Unit at Addenbrooke’s Hospital for the technical support in MRI. We acknowledge the help of Gaspar Delso (GE Healthcare) and Dattesh Shanbhag (GE Global Research) for their MRI motion correction programming code and Dimitris Voukantsis for his advice on data visualization.

