## Supplemental Material for "Multiparametric MRI of Early Tumor Response to Immune Checkpoint Blockade in Metastatic Melanoma"

**Table S1.** Patient demographics.

| Patient | ECOG | BRAF Status | Treatment | Standard CT Evaluation | | Body Site(s) Imaged | No. of Target Lesions Imaged | MRI Scan Completion | | |
|---|---|---|---|---|---|---|---|---|---|---|
| | | | | 12Weeks | 12 Months | | | Baseline | 3-Weeks | 12-Weeks |
| P1 | 1 | WT | Pembrolizumab | Progressive Disease | Progressive Disease | Subcutaneous | 2 | Yes | Yes | Yes |
| P2 | 1 | WT | Pembrolizumab | Progressive Disease | Progressive Disease | Liver | 1 | Yes | Withdrew from study (claustrophobia and disease progression) | |
| P3 | 0 | Mutant | Pembrolizumab | Partial Response | Partial Response | Inguinal and external iliac nodes | 2 | Yes | Yes | Yes |
| P4 | 1 | WT | Nivolumab | Mixed Response | Partial Response | Subcutaneous | 4 | Yes | Yes | Yes |
| P5 | 1 | WT | Nivolumab | Progressive Disease | Progressive Disease | Right occipital and posterior cervical region in the neck | 1 | Yes | Withdrew from study (disease progression) | |
| P6 | 1 | WT | Pembrolizumab | Progressive Disease | Progressive Disease | Lower limb | 1 | Yes | Yes | Withdrew from study (disease progression) |
| P7 | 0 | WT | Pembrolizumab | Mixed Response | Progressive Disease | Inguinal and external iliac nodes | 2 | Yes | Yes | Yes |

| | | | | | | | | | | |
|---|---|---|---|---|---|---|---|---|---|---|
| P8 | 0 | WT | Pembrolizumab | Mixed Response | Progressive Disease | Subcarinal/Paraesophageal nodes | 2 | Yes | Yes | Yes |
| P9 | 0 | WT | Combined Ipilimumab and Nivolumab | Partial Response | Partial Response | Cervical lymph node and supraclavicular | 2 | Yes | Yes | Yes |
| P10 | 0 | WT | Combined Ipilimumab and Nivolumab | Partial Response | Partial Response | Subcutaneous | 2 | Yes | Yes | Yes |
| P11 | 0 | Mutant | Combined Ipilimumab and Nivolumab | Progressive Disease | Progressive Disease | Medial clavicle | 1 | Yes | Withdrew from study (operation and disease progression) | |
| P12 | 0 | WT | Combined Ipilimumab and Nivolumab | Partial Response | Partial Response | Liver | 1 | Yes | Yes | Yes |
| P13 | 0 | WT | Combined Ipilimumab and Nivolumab | Partial Response | Partial Response | Adrenal and liver | 1 | Yes | Yes | Yes |
| P14 | 0 | Mutant | Combined Ipilimumab and Nivolumab | Partial Response | Partial Response | Peritoneal and mesenteric nodes | 3 | Yes | Yes | Yes |
| P15 | 1 | WT | Combined Ipilimumab and Nivolumab | Mixed Response | Progressive Disease | Inguinal and iliac nodes | 2 | Yes | Yes | Study halted due to COVID-19 crisis from March 2020 |

**Table S2.** MRI sequence parameters.

| Parameter | $T_2W$ | DKI | DCE |
|---|---|---|---|
| Sequence | SSFSE | 2D DW EPI | 3D FSPGR |
| TR (ms) | 1073-3231 | 4000-6667 | 3.2-3.3 |
| TE (ms) | 87.6-90.6 | 92.3-95.3 | 1.2-1.3 |
| Flip angle (º) | 90 | 90 | 16 |
| Slice thickness (mm) | 6 | 6 | 5 |
| Slice gap (mm) | 0 | 0 | 0 |
| FOV (cm) | 28-36 | 28-36 | 30-35 |
| Image matrix | 256 x 256 | 128 x 128 | 160 x 160 x N |
| Fractional k-space coverage | 0.54 | 1 | 0.52-0.73 |
| Parallel imaging factor | - | 2 | 2.5 |
| Acquisition time (min) | 3:39 | 11:30 | 8:07 |

*$T_2W$ = $T_2$-weighted anatomical scans; DKI = diffusion kurtosis imaging; DCE = dynamic contrast-enhanced MRI; SSFSE = single-shot fast spin-echo; 2D DW EPI = two-dimensional diffusion-weighted echo planar imaging; 3D FSPGR = three-dimensional fast spoiled gradient-recalled echo; TR = repetition time; TE = echo time; FOV = field of view*

Figure S1

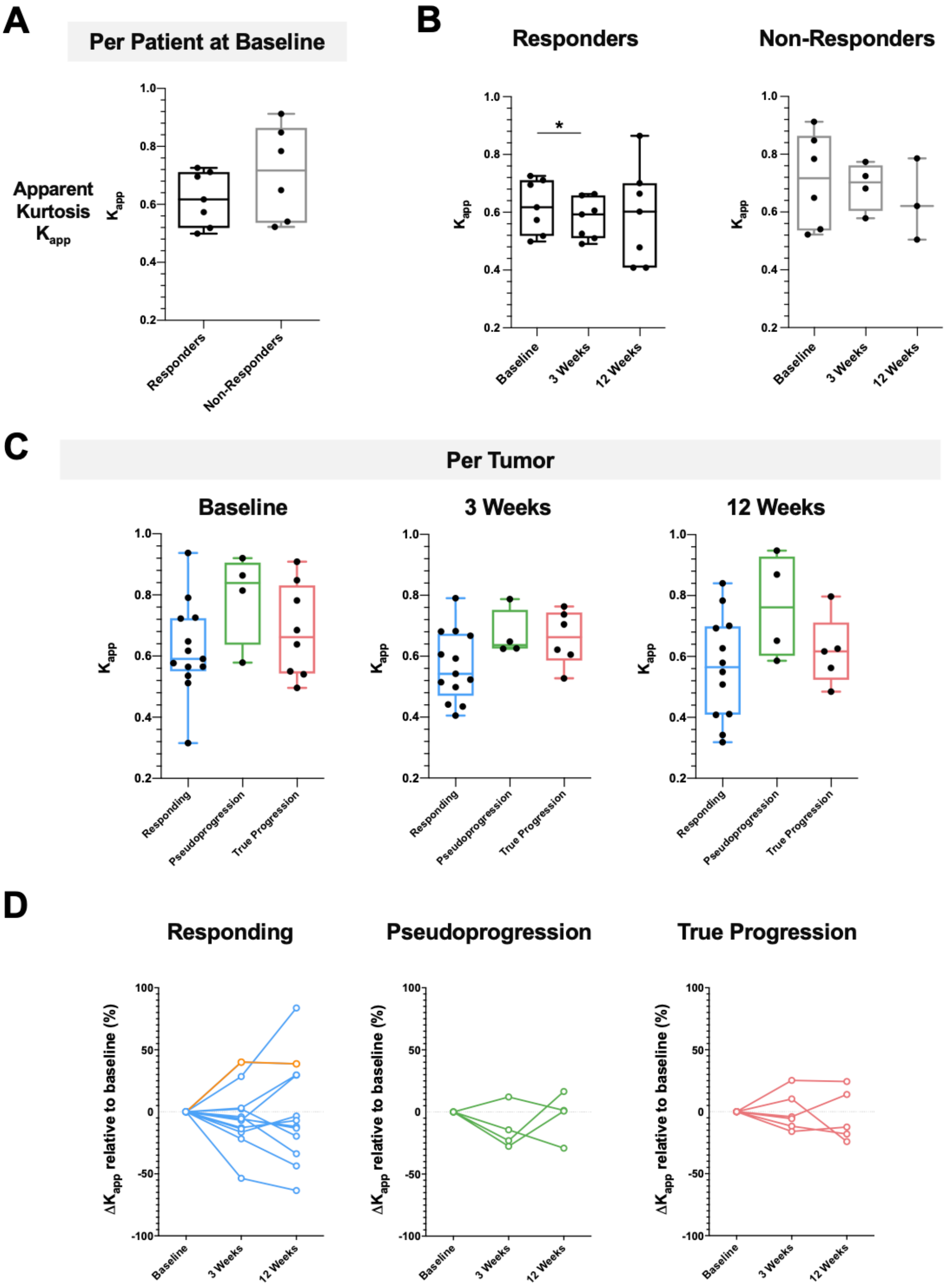

A
Per Patient at Baseline
Apparent Kurtosis $K_{app}$
$K_{app}$
Responders
Non-Responders
B
Responders
Non-Responders
Baseline
3 Weeks
12 Weeks
C
Per Tumor
Baseline
3 Weeks
12 Weeks
Responding
Pseudoprogression
True Progression
D
Responding
Pseudoprogression
True Progression
$\Delta K_{app}$ relative to baseline (%)

**Figure S1**

Measurement of tumor heterogeneity using DKI. (**A**) Comparison of apparent kurtosis ($K_{app}$) as a measure of tumor heterogeneity between responders and non-responders at baseline before the start of treatment. (**B**) Changes in tumor $K_{app}$ among the patients over the course of treatment. (**C**) Differences in tumor heterogeneity among the three subgroups of individual lesions at baseline. (**D**) Percentage change in $D_{app}$ relative to baseline in individual lesions from the three subgroups. * $p < 0.05$. Yellow line in (D) indicates the percentage change in $K_{app}$ for patient P4.

**Figure S2**

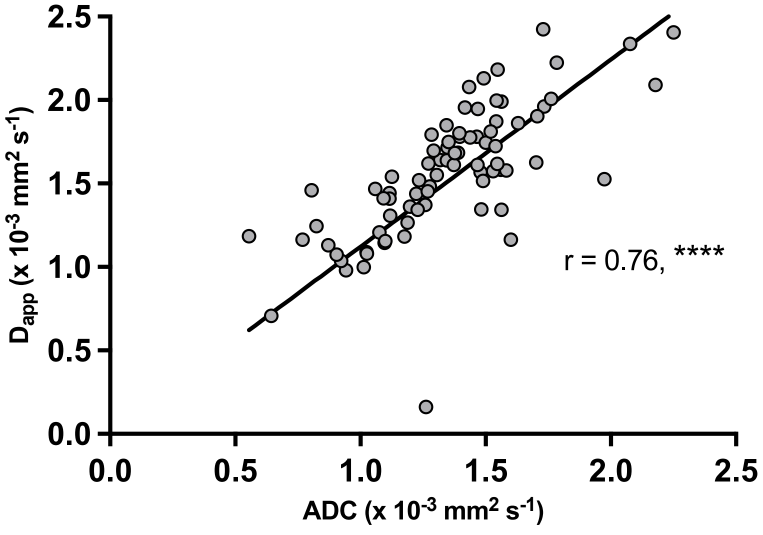


Spearman's correlation analysis showed a positive correlation between the apparent diffusivity value $D_{app}$ measured on DKI and apparent diffusion coefficient ADC measured on DWI. **** $p < 0.0001$.

**Figure S3**

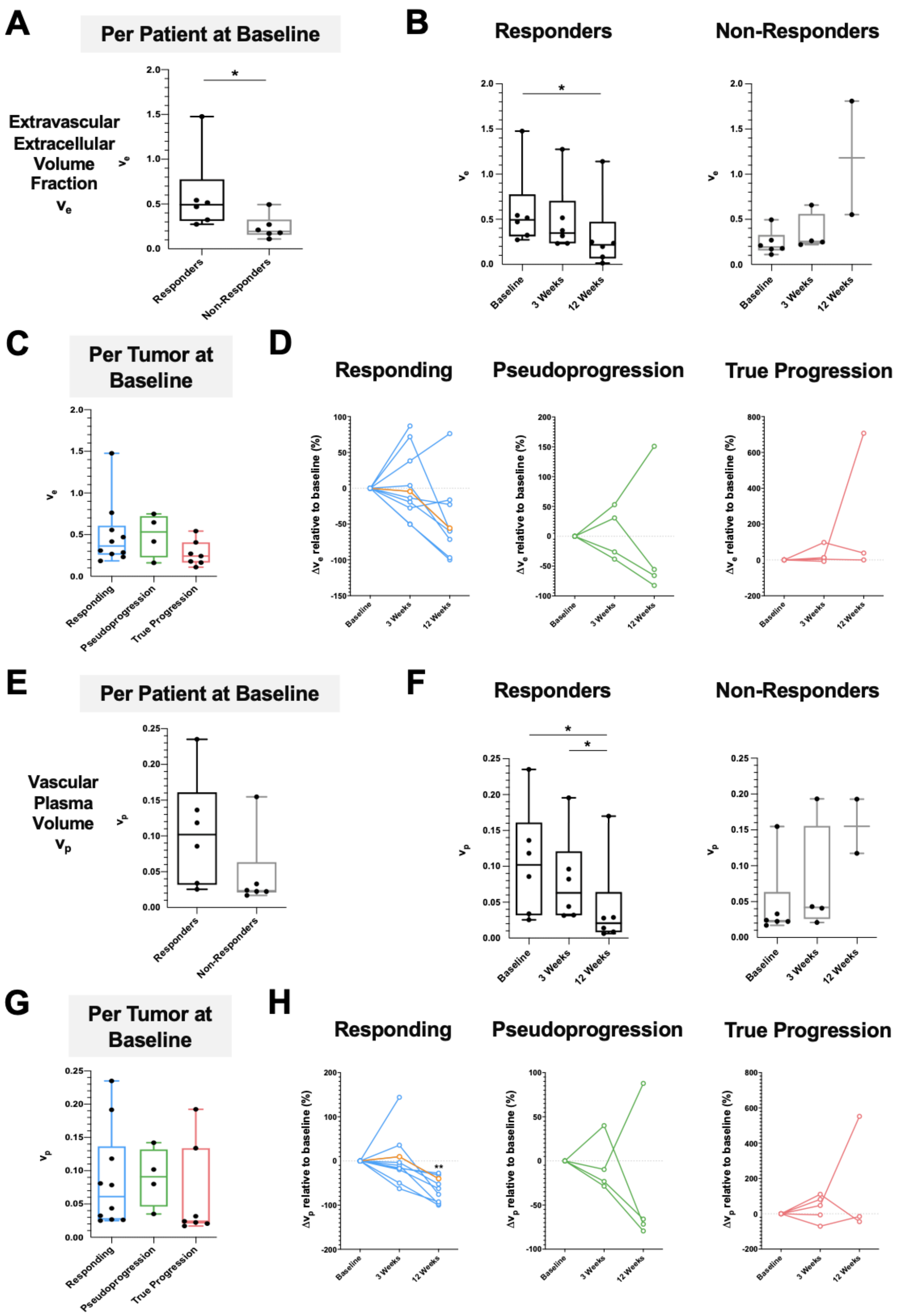

A
Per Patient at Baseline
Extravascular Extracellular Volume Fraction $v_e$
Responders
Non-Responders
B
Responders
Non-Responders
Baseline
3 Weeks
12 Weeks
C
Per Tumor at Baseline
Responding
Pseudoprogression
True Progression
D
Responding
Pseudoprogression
True Progression
$\Delta v_e$ relative to baseline (%)
E
Per Patient at Baseline
Vascular Plasma Volume $v_p$
F
Responders
Non-Responders
G
Per Tumor at Baseline
H
Responding
Pseudoprogression
True Progression
$\Delta v_p$ relative to baseline (%)

**Figure S3**

Comparison of changes in DCE-MRI parameters following treatment with immune checkpoint inhibitors among patients and within individual lesions: $v_e$, extravascular extracellular volume fraction (A-D); and $v_p$, vascular plasma volume (E-H). * $p < 0.05$; ** $p < 0.01$. Yellow line in (D) and (H) indicates the percentage change in $v_e$ and $v_p$ for patient P4.

**Figure S4**

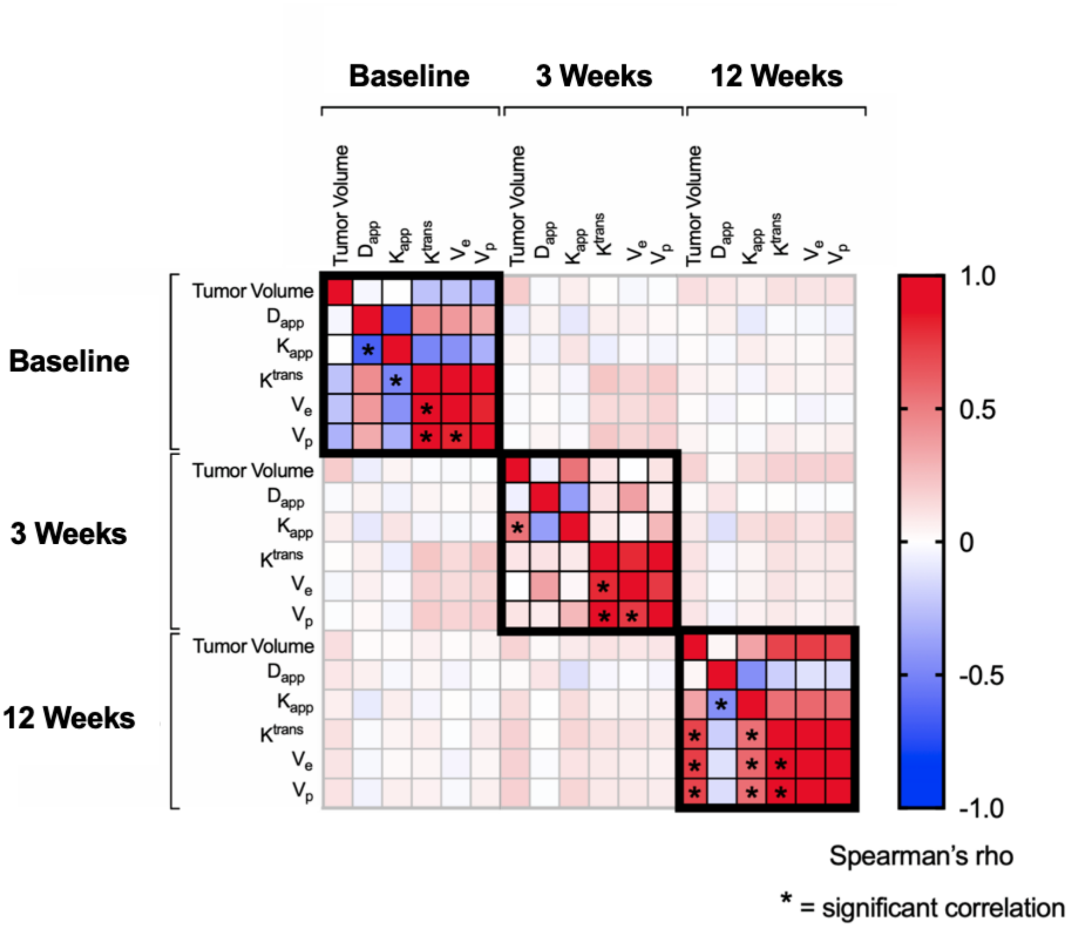


Spearman's correlation analysis of the MRI imaging biomarkers during the first 12 weeks of immune checkpoint blockade.

### Supplemental Methods

#### MRI Image Acquisition

$T_2$-weighted SSFSE images were acquired in coronal, axial, and sagittal orientations (TE/TR = 90.6/1508 ms; slice thickness/gap 6.0/0.0 mm; matrix 256 x 256, NEX 0.54). Axial DKI was acquired using dual-echo EPI (TE/TR = 92.3/6000 ms; slice thickness/gap 6.0/0.0 mm; matrix 128 x 128) with five different $b$-values (100, 500, 900, 1300 and 1700 s/mm$^2$). A $b$-value of 0 s/mm$^2$ was collected but not used as low $b$-values have been reported to be susceptible to pseudo-perfusion bias that can be avoided using $b$-value >100 s/mm$^2$. Standard diffusion-weighted imaging (DWI) was performed using $b$-values of 100, 500, 900 s/mm$^2$. DCE-MRI was performed using a three-dimensional segmented k-space spoiled gradient-echo (3D SPGR) technique in a sagittal orientation. Dynamic phases were acquired at a temporal resolution of 12 s, which included approximately 7 s of scanning and a 5 s gap. The subjects were trained to hold their breath during the gradient noise and to breathe in and out during the periods of silence (TR/TE = 1.2/3.2 ms; flip angle 16$^o$; slice thickness/gap 5.0/0.0 mm; matrix 160 x 160 x number of slices). A bolus injection of the intravascular contrast agent Gadobutrol (Gadovist, Bayer Pharma AG, Berlin, Germany) was administered at 0.1 mmol/kg intravenously 25 s after the start of acquisition at a rate of 3 ml/s, followed by 25 ml of saline flush using an MR-compatible power injector. Pre-contrast $T_1$-weighted gradient-echo images were acquired with five different flip angles (2$^o$, 3$^o$, 5$^o$, 10$^o$, 15$^o$) and Bloch-Siegert $B_1$ maps[1] were obtained for subsequent $B_1$ inhomogeneity correction and $T_1$ mapping for analysis of the dynamic sequences.

#### Image Processing and Analysis

Quantitative maps of $D_{app}$ were calculated from the DKI images using an in-house Matlab Script (Mathworks, MA, USA). Non-Gaussian water movements were quantified using a dimensionless metric termed kurtosis, governed by the polynomial equation (**Equation 1**):

$$S_i = S_0 \cdot \exp\left(-b_i \cdot D_{app}\right) \cdot \exp\left(\frac{1}{6} \cdot b_i^2 \cdot {D_{app}}^2 \, . \, K_{app}\right) \qquad (1)$$

where $s_i$ is the signal intensity at b-value $b_i$, $s_0$ is the estimated signal intensity at $b$-value $b_i$ = 0 s/mm$^2$ when no diffusion gradient is applied, $D_{app}$ is the apparent diffusivity (s/mm$^2$) at $b$-values more than 1000 s/mm$^2$, and $K_{app}$ is the apparent diffusion kurtosis (unitless)[2,3].

Quantitative maps of tumor vascular permeability and perfusion were calculated from the DCE images using MISTAR software (Apollo Medical Imaging Technology, Melbourne, Australia). $B_1$ maps of field inhomogeneity were calculated using in-house Matlab scripts for subsequent $B_1$-correction of pre-contrast $T_1$ maps. A model arterial input function (AIF) based on Fritz-Hansen *et al.*[4] and the extended Tofts model[5,6] were used for pharmacokinetic modelling of the intravascular contrast enhancement within tissues. Total tissue contrast concentration as a function of time $[C_e(t)]$ can be approximated using (**Equation 2**):

$$C_e(t) = v_p C_p(t) + K^{trans} \int_0^t C_p(t)\, e^{-k_{ep}(t-\tau)}\, d\tau \tag{2}$$

where $C_p(t)$ is the concentration in blood plasma as a function of time as determined by the AIF measured in a large feeding vessel, $K^{trans}$ is the volume transfer coefficient from the blood plasma space into the extravascular tumor interstitial space reflecting vascular permeability, $v_p$ is the vascular plasma volume, $v_e$ is the fractional volume of the extravascular extracellular space, and $k_{ep}$ is the flux rate constant ($K^{trans} = v_e \times k_{ep}$). Motion correction was performed on all images using in-house Matlab scripts prior to the calculation of quantitative maps.

Volumetric tumor regions of interest (VOIs) were drawn on the $T_2$-weighted images and quantitative maps of $D_{app}$, $K_{app}$, $K^{trans}$ $v_e$ and $v_p$ by a radiologist (FS) with 3 years of experience in the oncological setting and blinded to the clinical outcome and a radiology researcher (DL) with 4 years of experience in oncological imaging. VOIs from each tumor were expressed as single $T_2$ volume, $D_{app}$, $K_{app}$, $K^{trans}$ $v_e$ and $v_p$ values and volume-weighted for analysis for the patient as a whole or for individual lesion. $T_2$-weighted images taken at each imaging timepoints were normalized based on intensity scaling to the baseline images before calculating the $T_2$ histograms[7].

**Immunohistochemistry**

All immunochemistry (IHC) procedures were performed on histologically confirmed melanoma tissues obtained for diagnosis. Tissues were fixed with 10% buffered formalin solution and processed into paraffin embedded (FFPE) tissue blocks. Serial tissue sections of 3.5 µm thickness were prepared from the FFPE blocks and mounted onto silanized slides at

Addenbrooke's Hospital Human Research Tissue Bank. IHC and semi-automated image analysis were carried out at AstraZeneca UK. Slides were baked for 1 h at 60$^{o}$C, deparaffinized in xylene and rehydrated with decreasing concentrations of ethanol. Automated H&E staining was performed on Leica ST5020 Multistainer (Leica Biosystems, Germany). Immunostaining for CD31, Ki67, CAIX, CD8, FOXP3 and CD11b was conducted on a Ventana Benchmark ULTRA automated slide processing system (Ventana Medical Systems Inc., Roche Tissue Diagnostics, Germany) according to the manufacturer's instructions. Briefly, heat-induced epitope retrieval in Tris-EDTA buffer pH 7.8 at 95$^{o}$C for 44 min was carried out on all slides using Ventana's ULTRA Cell Conditioning 1 (CC1) solution, and endogenous peroxidase activity was quenched using DISCOVERY Inhibitor (Roche Tissue Diagnostics, Germany) at 37$^{o}$C for 4 min. Primary antibodies incubation was conducted at 37$^{o}$C for 1 h using the following monoclonal anti-human antibodies: mouse anti-human CD31 (1 μg/ml, clone JC70A, Dako, Denmark), CONFIRM$^{TM}$ rabbit anti-human Ki67 (2 μg/ml, clone 30-9, Roche Tissue Diagnostics, Germany), rabbit anti-human carbonic anhydrase IX (CAIX) (2 μg/ml, clone EP161, Roche Tissue Diagnostics, Germany), rabbit anti-human CD8 (0.58 μg/ml, clone SP239, Spring Bioscience, USA), rabbit anti-human Foxp3 (0.20 μg/ml, clone SP97, Spring Bioscience, USA) and rabbit anti-human CD11b (0.03 μg/ml, clone D6X1N, Cell Signaling Technology, USA). This was followed by secondary antibodies incubation at 37$^{o}$C for 16 min using either DISCOVERY$^{TM}$ high quality horseradish peroxidase (HQ-HRP) kit for highest sensitivity and specificity in detection and chromogen staining using EnVision FLEX HRP Magenta Substrate Chromogen System (Dako, Denmark), DISCOVERY$^{TM}$ Purple HRP-activated Chromogen kit or DISCOVERY$^{TM}$ Teal HRP-activated Chromogen kits (Roche Tissue Diagnostics, Germany). Human tonsil, spleen and placenta were used as positive control tissues. Immunostaining with mouse or rabbit negative isotype control antibodies (Roche Tissue Diagnostics, Germany) was used as negative controls. Slides were then counterstained with hematoxylin. All stained slides were scanned at 20X and 40X magnification on Aperio AT2 (Leica Biosystems, Germany). Semi-automated image analysis of the whole tissue slides was performed on HALO$^{®}$ software (Indica Labs, USA).
